# Novel Tools for Lassa Virus Surveillance in Peri-domestic Rodents

**DOI:** 10.1101/2023.03.17.23287380

**Authors:** Allison R. Smither, James Koninga, Franklyn B. Kanneh, Momoh Foday, Matthew L. Boisen, Nell G. Bond, Mambu Momoh, John Demby Sandi, Lansana Kanneh, Foday Alhasan, Ibrahim Mustapha Kanneh, Mohamed S. Yillah, Donald S. Grant, Duane J. Bush, Diana K. S. Nelson, Kaitlin M. Cruz, Raphaëlle Klitting, Matthias Pauthner, Kristian G. Andersen, Jeffrey G. Shaffer, Robert W. Cross, John S. Schieffelin, Robert F. Garry

## Abstract

**Background:** Lassa fever (LF) is a rodent-borne disease endemic to West Africa. In the absence of licensed therapeutics or vaccines, rodent exclusion from living spaces remains the primary method of preventing LF. Zoonotic surveillance of Lassa virus (LASV), the etiologic agent of LF, can assess the burden of LASV in a region and guide public health measures against LF.

**Methods:** In this study, we adapted commercially available LASV human diagnostics to assess the prevalence of LASV in peri-domestic rodents in Eastern Sierra Leone. Small mammal trapping was conducted in Kenema district, Sierra Leone between November 2018-July 2019. LASV antigen was detected using a commercially available LASV NP antigen rapid diagnostic test. LASV IgG antibodies against LASV nucleoprotein (NP) and glycoprotein (GP) were tested by adapting a commercially available semi-quantitative enzyme linked immunosorbent assay (ELISA) for detection of mouse-related and rat-related species IgG.

**Findings:** Of the 373 tested specimens, 74 (20%) tested positive for LASV antigen. 40 (11%) specimens tested positive for LASV NP IgG, while an additional 12 (3%) specimens only tested positive for LASV GP IgG. Simultaneous antigen presence and IgG antibody presence was linked in *Mastomys sp*. specimens (*p* < 0.01), but not *Rattus sp*. specimens (*p* = 1). Despite the link between antigen presence and IgG antibody presence in *Mastomys sp*., the strength of antigen response did not correlate with the strength of IgG response to either GP IgG or NP IgG.

**Interpretation:** The tools developed in this study can aid in the generation of valuable public health data for rapid field assessment of LASV burden during outbreak investigations and general LASV surveillance.

**Funding:** Funding for this work was supported by the National Institute of Allergy and Infectious Diseases National Institute of Health, Department of Health and Human Services under the following grants: International Collaboration in Infectious Disease Research on Lassa fever and Ebola - ICIDR - U19 AI115589, Consortium for Viral Systems Biology - CViSB - 5U19AI135995, West African Emerging Infectious Disease Research Center - WARN-ID - U01AI151812, West African Center for Emerging Infectious Diseases: U01AI151801.

## Introduction

Lassa mammarenavirus (LASV) is an important public health threat for much of West Africa, where approximately 100,000-300,000 cases of Lassa fever occur annually with an estimated 5,000 deaths.^1^ LASV is a bi-segmented ambisense RNA virus and encoding of four multifunctional proteins: the nucleoprotein (NP), the RNA-dependent RNA polymerase large, a matrix RING-finger Zinc-binding protein, and the glycoprotein complex (GP).^2^ A rodent-borne pathogen, LASV is primarily transmitted from infected rodents to humans via inhalation or ingestion of rodent excreta, though human-human and nosocomial transmission accounts for approximately 20% of Lassa fever cases.^3^ With no licensed therapeutics or vaccines currently available for Lassa fever, rodent exclusion and pest management programs are recommended for prevention of LASV infection.

The primary host reservoir associated with LASV is the multimammate mouse *Mastomys natalensis*. In recent years, two other closely related species, *Mastomys erythroleucus* and *Hylomyscus pamfi*, have been identified as additional host reservoirs of LASV in Nigeria and Guinea.^4^ LASV Immunoglobulin G (IgG) has also been detected in other common rodent species such as *Rattus rattus* and *Mus musculus* in specimens collected from LASV-endemic areas.^5-7^ *Mastomys, Hylomyscus, Rattus*, and *Mus* genus are all broadly distributed throughout sub-Saharan Africa; yet *M. natalensis* is the most widely distributed rodent species on the continent. Despite the near-ubiquitous presence of *M. natalensis* in sub-Saharan Africa, LASV is known to only circulate in West Africa. Nonetheless, climate change alongside changes in land use and increases in population density will likely allow for expansion of the ecological niche of LASV and the risk of Lassa fever further into Central and Eastern Africa in coming decades.^8^ Detection of active and previous LASV infections in the rodent hosts provides crucial knowledge for public health assessments of LASV risk in both LASV-endemic regions and regions at risk of LASV expansion.

Numerous enzyme-linked immunosorbent assays (ELISA) have been developed to detect LASV antigen and antibody.^9-11^ Rapid diagnostic tests (RDTs) have also been created for LASV antigen with similar concordance to antigen ELISA.^9,12^ Unlike ELISA which must be performed in a laboratory setting, an RDT can be performed in low-resource settings and give results in as little as 15 minutes. The ability to detect a potentially active LASV infection in a rodent at the point of collection could allow for immediate implementation of public health interventions in the community of sampling, reducing risk of contracting LASSA FEVER. Meanwhile, rodent surveillance for LASV IgG has traditionally been performed using indirect immunofluorescence assay (IFA).^6^ However, IFA require Biosafety Level 4 (BSL-4) facilities to perform, making it impractical for in-country surveillance. Recently, Soubrier and colleagues adapted a human LASV IgG ELISA kit for detection of LASV IgG in *Mastomys sp*. using anti-mouse (*Mus*) reagents, with similar concordance to IFA.^13^ However, this assay only 1) qualitatively indicates presence or absence of IgG, as opposed to the strength of IgG response 2) detects IgG against *Mus*-related species and 3) measures only NP IgG.

LASV NP is the most abundantly expressed and immunogenic of the four LASV proteins. NP is understood to dampen the innate immune response by suppressing double-stranded RNA; antibodies to NP do not exhibit neutralizing activity.^14^ Conversely, LASV glycoprotein (GP) IgG antibodies are crucial for antibody-mediated neutralization of LASV as GP is the sole antigen displayed on the surface of the virion.^15^ Although an ELISA cannot determine neutralizing activity of an antibody, the strength of a potentially neutralizing GP IgG response compared to the strength of the predominant NP IgG response can provide insight into routes and duration of natural LASV infection and prioritize samples for further neutralization assay testing. Furthermore, it has been demonstrated that neonatal rodents infected with LASV or other mammarenaviruses develop persistent infections and continually shed virus and antigen despite low-level presence of antiviral antibodies.^16,17^ *Mastomys sp*. infected with mammarenaviruses as adults, on the other hand, rapidly clear the virus and develop high levels of antiviral antibodies.^18,19^ Understanding the link between concurrent antigen and antibody presence could also help to further elucidate the ecological niches permissive to LASV infection.

The goal of this study was to adapt commercially available LASV human diagnostics for rodent sero-surveillance of LASV to assess the prevalence of LASV in peri-domestic rodents in Eastern Sierra Leone. Rodent sero-surveillance data in Sierra Leone is limited, with only one study published since the 1991-2002 Sierra Leone Civil War.^20,21^ We assessed the presence of LASV NP in peri-domestic rodents using a field-deployed antigen rapid test to quickly determine prevalence of LASV antigen in the study population. Next, we modified a human semi-quantitative IgG ELISA for LASV to detect NP and GP IgG in both *Mastomys*-related and *Rattus*-related species. Lastly, we examined the relationship of the NP and GP IgG responses and the relationship between antigen and IgG presence among species. Together, these tools and data help to provide updated estimates of the prevalence of LASV in Eastern Sierra Leone rodents.

## Materials and methods

### Specimen collection and processing

Small mammal trapping was conducted in Kenema District, Eastern Province, Sierra Leone between November 2018-July 2019. Both the Sierra Leone Ethics and Scientific Research Committee and Tulane University Institutional Animal Care and Use Committees reviewed and approved this study (Protocol ID 1034). Rodent trapping and processing protocols were followed as previously described.^22^

### Antigen rapid diagnostic test (RDT)

The ReLASV Pan-Lassa Antigen Rapid Test (RDT) (Zalgen, MD, USA) was deployed and scored during field collections in accordance with the manufacturer’s instructions using 30 μL of fresh rodent serum. Presence of LASV antigen detected by RDT has been shown to strongly correlate with antigen detected by ELISA.^9^

### Rodent species identification

Identification to the genus level was performed in the field. Prior to IgG ELISA, specimens were classified as either mouse-related specimens *(Mastomys sp*., *Praomys sp*., and *Hylomyscus sp*.) or rat-related specimens (*Rattus sp*.) to ensure the appropriate ELISA controls and secondary antibodies were utilized for the respective specimens. Cytochrome *b* polymerase chain reaction (PCR) of DNA extracted from tissue samples was used to confirm specimen identity to the species level as previously described.^23^

### Enzyme-Linked Immunosorbent Assay (ELISA)

All ELISAs were performed using Zalgen ReLASV Pan-LASV Pre-fusion (Pf) -GP and NP IgG kits as previously described, with adaptations to allow for detection of rodent antibodies.^24^ Horseradish peroxidase-labeled (HRP) secondary antibody solutions (Donkey anti-Rat IgG (H+L) HRP [Thermofisher, CA, USA] diluted 1:2000 in Zalgen Undyed Antibody Conjugate; Goat anti-Mouse IgG(γ), HRP [Seracare, MA, USA] diluted 1:2500 in Zalgen Undyed Antibody Conjugate Diluent) were used in place of the anti-human secondary antibody solution provided in the kits. Mouse and rat positive and negative controls were used in place of the provided lyophilized human controls. All other kit components, including the antigen coated ELISA plates, sample diluent, wash solution (PBS-Tween 20), 3,3’,5,5’-Tetramethylbenzidine (TMB) substrate, and stop solution (2% methanesulfonic acid) from the kits were used in accordance with the manufacturer’s instructions. Details of the adapted positive and negative controls, ELISA operation, and establishment of assay cutoffs are described in supplemental information.

### Statistical analysis

Unpaired two-tailed Student’s t-tests were used to compare the mean concentration between NP and GP IgG, respectively, in *Mastomys sp*. and *Rattus sp*. Correlations between IgG concentration from *Mastomys sp*. specimens positive for both GP IgG and NP IgG and mean IgG concentration versus antigen strength by protein were independently assessed by linear regression. Fisher’s exact test was used to compare proportions of antigen-positive and IgG-positive specimens. The raw data, compiled dataset and scripts used to generate the tables and figures are available at github.com/allisonrs/Project_LASV_AgAb_Rodent. A further description of programs and files used for statistical analysis are detailed in supplemental information.

## Results

### Study Population

A total of 373 rodent specimens were tested out of 534 small mammal specimens collected in 23 villages during sample collection (Figure 1, Table S1). We excluded specimens (N = 161) that did not yield enough serum volume to perform both antigen and antibody testing, as well as specimens that were not closely related to either *Mus* or *Rattus* species (i.e. *Crocidura sp*.). For in-country ELISA testing, specimens were classified to the genus level based off morphometric features in the field. Select specimens were confirmed to the species level by Cytochrome *b* PCR (Table S2). All *Mastomys sp*. tested were identified as *M. natalensis*, all *Rattus sp*. tested were identified as *R. rattus*. In addition, the *Praomys sp*. specimens tested were identified as *P. rostarus* and the single *Hylomyscus sp*. specimen was identified as *H. simius*. The single field identified *Malacomys sp*. specimen was later identified as *P. rostarus* by Cytochrome *b* PCR.

**Figure 1:**
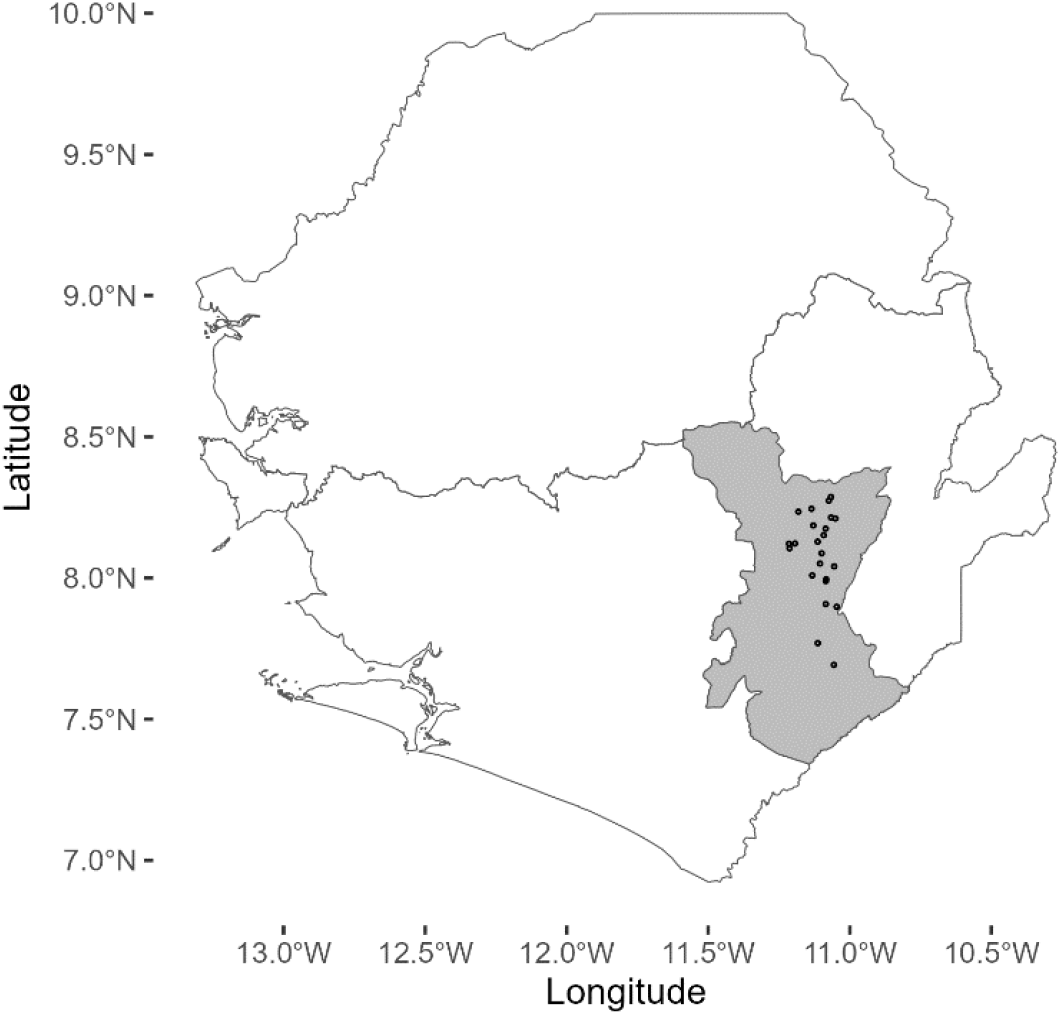
Provincial level map of Sierra Leone with location of villages trapped as part of study. Each village is represented by a dot. The Kenema district is shaded in gray.

### In-field LASV NP Antigen RDT and LASV NP IgG ELISA

LASV NP antigen was detected at the time of necropsy using the Zalgen ReLASV Pan-Lassa Antigen Rapid Diagnostic Test. A total of 74 of the 373 specimens (20%) tested positive for antigen (Table 1). 70 of the 74 antigen positive specimens (95% of the antigen positive population) were *Mastomys sp*. 3 (4 %) *Rattus sp*. tested antigen positive, and one (<1%) *Praomys sp*. tested antigen positive. For the NP IgG ELISA, 234 specimens (*Mastomys, Praomys, Hylomyscus sp*.) were tested with anti-mouse reagents, and 139 specimens (*Rattus sp*.) were tested with anti-rat reagents (Table S3). A total of 40 specimens (11%) tested positive for NP IgG (Table S4). 36 of the 40 NP IgG positive specimens (90% of the NP IgG positive population) were *Mastomys sp*., and four (10%) were *Rattus sp*. No *Praomys sp*. or *Hylomyscus sp*. tested positive for NP IgG.

**Table 1:** In-field Antigen RDT results by genus with row percentages. *Includes 1 field identified *Malacomys* specimen later confirmed to be *P. rostratus* by cytochrome *b* PCR.

| **Antigen Positive Specimens by Genus** |  |  |
| --- | --- | --- |
| **Genus** | **N** | **Ag+ (% of Genus)** |
| *Mastomys sp.* | 219 | 70 (32) |
| *Rattus sp.* | 139 | 3 (2) |
| *Praomys sp.* * | 12 | 1 (8) |
| *Hylomyscus sp.* | 3 | 0 (0) |
| Total | 373 | 74 (20) |

### Comparison between NP IgG and GP IgG Response

We next wanted to compare the NP IgG response to the GP IgG response in mouse-related species versus the rat-related species. An additional 12 specimens tested positive exclusively for GP IgG, for a total of 52 specimens (16% of the total population) IgG positive to at least one protein (Table 2). Interestingly, an equal number of *Rattus sp*. and *Mastomys sp*. were GP IgG positive only. Among the specimens that tested positive for both GP IgG and NP IgG, 21 (91%) of the 23 GP+/NP+ had higher NP IgG concentrations than GP IgG concentrations (Figure 2). The mean NP IgG concentration of the IgG+ *Mastomys sp*. population was higher than its mean GP IgG concentration (Figure 2, unpaired Student’s two-tailed t-test, *p* < 0.01). Indeed, linear regression analysis indicated NP IgG concentrations were weakly correlated with GP IgG concentrations in *Mastomys sp*. (R^2^ = 0.2463) (Figure S1). In contrast, seven of the ten (70%) IgG+ *Rattus sp*. specimens were only GP IgG+ or had a higher GP IgG concentration than NP IgG concentration. However, the difference in mean concentration between NP IgG and GP IgG was not statistically significant (Figure 2, unpaired Student’s two-tailed t-test, *p* = 0.69).

**Table 2:** GP and NP IgG ELISA results by genus with row percentages. *Includes 1 field identified *Malacomys sp*. specimen later confirmed to be *P. rostarus* by cytochrome b PCR.

| NP and GP IgG Positives by Genus |  |  |  |  |  |  |
| --- | --- | --- | --- | --- | --- | --- |
| Genus | N | IgG- | GP IgG+ only | NP IgG+ only | GP and NP IgG+ | Total IgG+ |
| *Mastomys sp.* | 219 | 177 (81) | 6 (3) | 14 (6) | 22 (10) | 42 (19) |
| *Rattus sp.* | 139 | 129 (93) | 6 (4) | 3 (2) | 1 (1) | 10 (7) |
| *Praomys sp.* * | 12 | 12 (100) | 0 (0) | 0 (0) | 0 (0) | 0 (0) |
| *Hylomyscus sp.* | 3 | 3 (100) | 0 (0) | 0 (0) | 0 (0) | 0 (0) |
| Total | 373 | 323 (86) | 12 (3) | 17 (5) | 23 (6) | 52 (14) |

**Figure 2:**
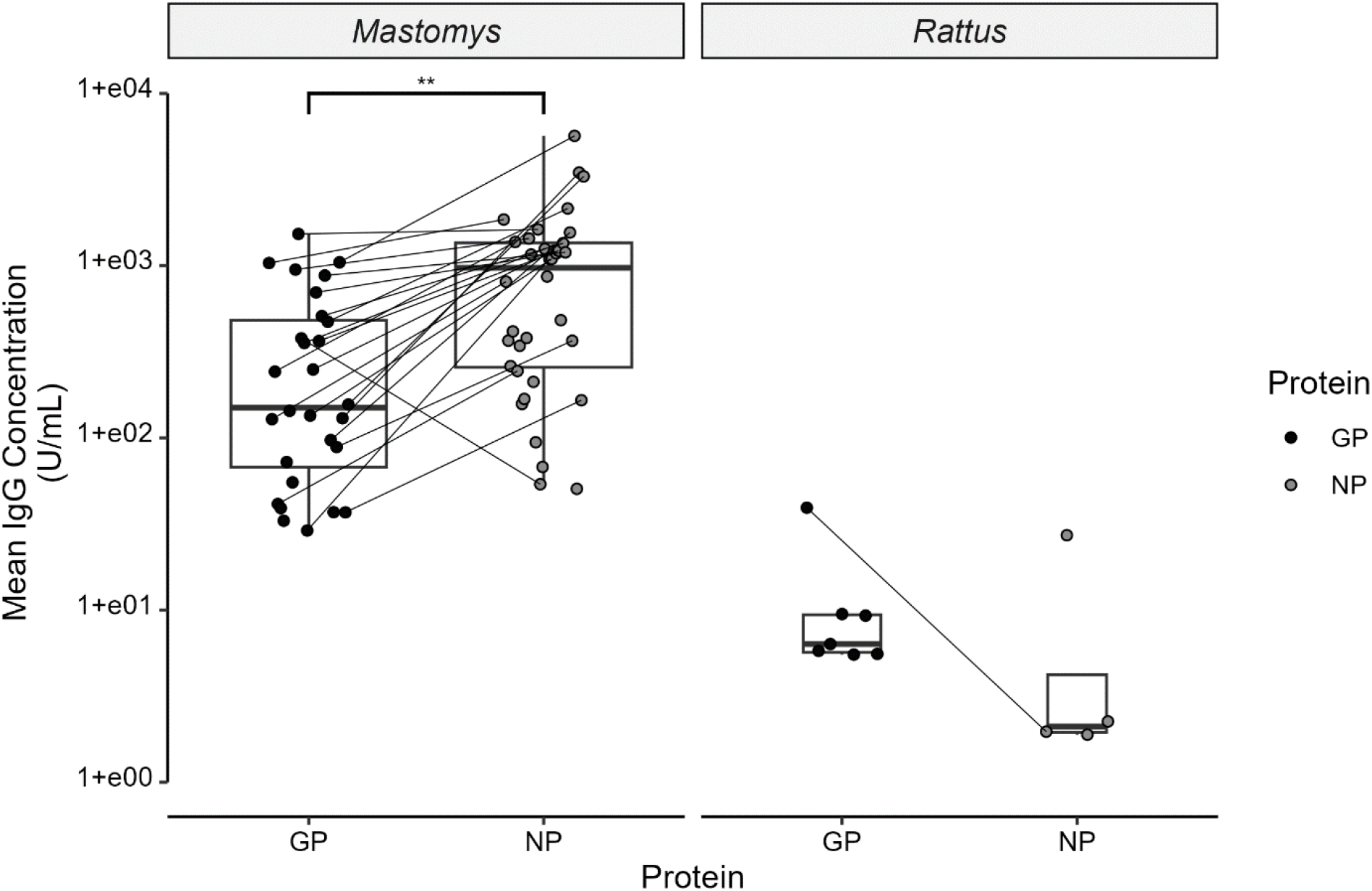
Distribution of positive GP IgG and NP IgG concentrations, grouped by genus. Specimens testing positive for both GP IgG and NP IgG are shown as connected pairs. Comparison of means performed by unpaired two-tailed Student’s t-test, ** = *p* < 0.01 (*Mastomys sp*.), *p* = 0.69 (*Rattus sp*.).

### Relationship between antigen and antibody presence

Simultaneous presence of LASV antigen and LASV IgG to at least one protein was linked in the *Mastomys sp*. population, where 21 of the 42 IgG+ specimens (50%) were also Ag+ (Table 3, *p* < 0.01 by Fisher’s Exact Test). Of the Ag+/IgG+ *Mastomys sp*. specimens, no correlation was observed between the amount of antigen present as indicated by RDT score and antibody concentration (Figure 3; GP IgG R^2^ = 0.0001, NP IgG R^2^ = 0.0032; both by linear regression). In contrast, none of the ten IgG+ *Rattus sp*. specimens tested Ag+ (Table S5, *p* = 1 by Fisher’s Exact Test).

**Table 3:** Contingency table of antigen RDT and IgG ELISA results for *Mastomys sp*.†p-values from Fisher’s exact test for comparing proportions of antigen (Ag) positive and IgG antibody positive specimens.

| Contingency Table of Antigen and Antibody Presence for *Mastomys sp.* |  |  |  |  |
| --- | --- | --- | --- | --- |
|  | IgG - | IgG + | Total | *p*-value† |
| Ag - | 128 | 21 | 149 | <0.01 |
| Ag + | 49 | 21 | 70 |  |
| Total | 177 | 42 | 219 |  |

**Figure 3:**
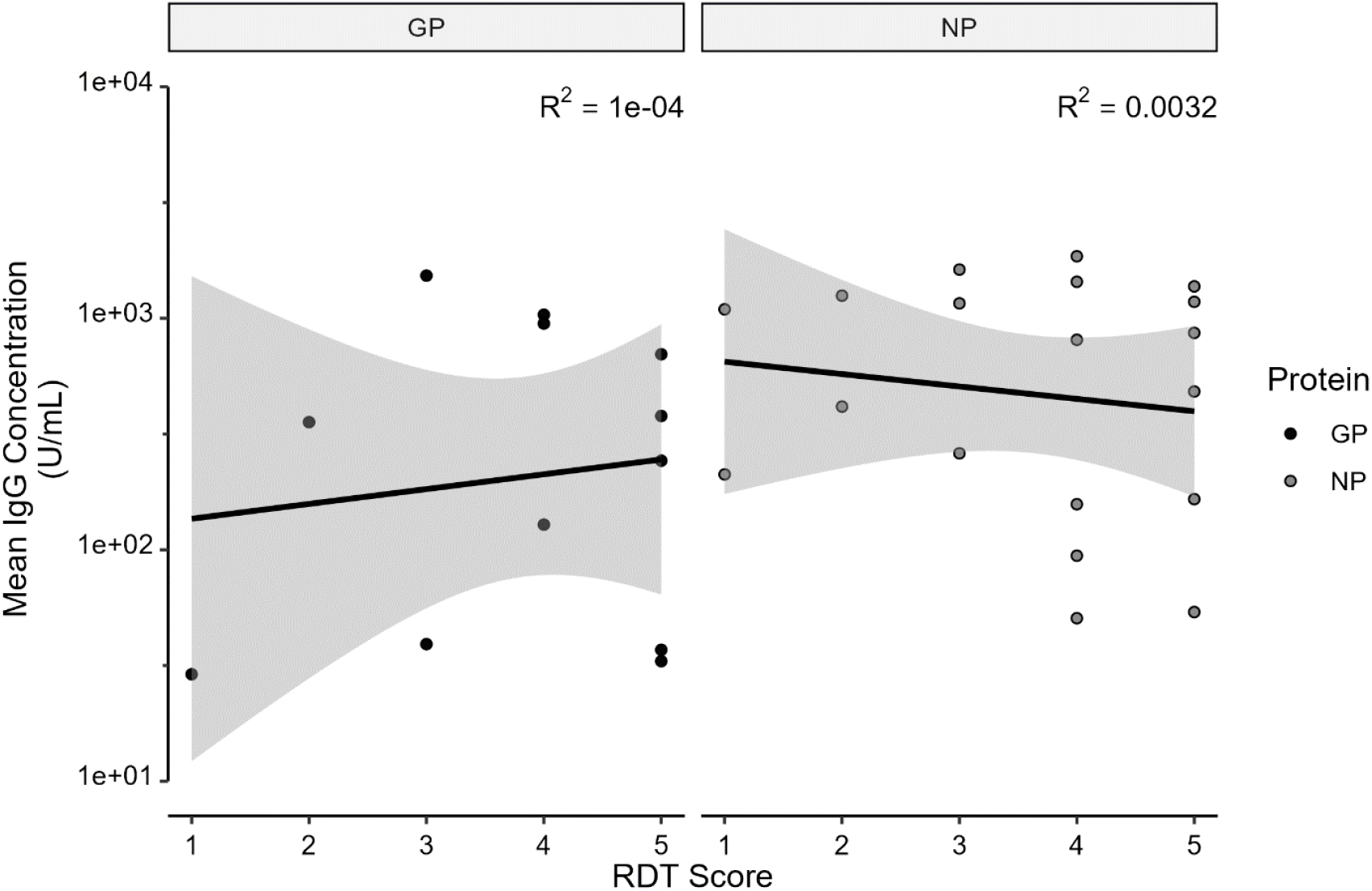
Mean IgG concentration vs. RDT score (antigen strength) of Ag+/IgG+ *Mastomys sp*., by antibody protein (Linear regression + standard error).

## Discussion

The proliferation of rapid diagnostic tests during the COVID-19 pandemic has solidified the utility of simple, on-demand, low-cost diagnostic tests in medical decision making. Their portability, ease of use and relative stability in a variety of temperatures also make RDTs ideal for field surveillance of pathogens. Likewise, we have demonstrated the ability to use commercially available LASV diagnostics for in-country rodent sero-surveillance. Our study is the first to use commercially available LASV antigen RDTs at the time of necropsy to quickly assess presence and relative strength of LASV antigen in collected rodents. Furthermore, whereas most studies surveying LASV IgG in rodents have only assessed presence or absence of LASV IgG via IFA, we developed a semi-quantitative ELISA to detect LASV IgG to both NP and GP in both mouse- and rat-related rodent species.

Of the 373 specimens examined in the study, 74 (20 % of study population) tested positive for LASV NP antigen (Table 1), in line with the single previous study to assess LASV antigen in peri-domestic rodents.^5^ Antigen was detected in *Mastomys sp*., *Rattus sp*., and *Praomys sp*.; LASV antigen has not been previously detected in *Praomys sp*. Of note, of LASV antigen tests (both RDT and ELISA) have been demonstrated to perform as well or better to detect active LASV infection compared with nucleic acid amplification tests.^9^ This is due to the substantial genetic diversity among LASV lineages and strains, while protein sequences among intra-lineage strains remain relatively conserved. Antigen presence alone, however, cannot be exclusively used to diagnose an active LASV infection.^25^ In the context of in-field surveillance, the RDT would be better situated as an initial diagnostic to triage specimens for further analyses, including nucleic acid tests or genomic sequencing.

After examining collected specimens for LASV NP antigen by RDT, we next examined the prevalence of LASV NP IgG following development of Mus-specific and Rattus-specific ELISA. 40 specimens (11% of study population) tested positive for NP IgG (Table S4), in line with other seroprevalence studies of similar scope and size.^5,6^ An additional 12 specimens tested IgG positive exclusively for LASV GP, for a total of 52 specimens (14% of the study population) IgG positive to at least one of the two proteins. Furthermore, six of the ten *Rattus sp*. specimens that were IgG positive were positive only to GP (Table 2). Only *Mastomys sp*. and *Rattus sp*. specimens tested IgG positive; we did not detect IgG in *Praomys sp*. or *Hylomyscus sp*. specimens (Table 2). However, LASV IgG has been previously detected in *Praomys sp*., specifically *P. daltoni*.^26^ In addition, *Hylomyscus pamfi* has been documented as a novel LASV reservoir in Nigeria (both IgG positive and real-time PCR positive) and as a host of a related LASV-like virus in Cote d’Ivoire.^7,27^ Given that these studies have used anti-mouse IgG secondary antibodies to detect LASV IgG in both *Praomys sp*. and *Hylomyscus sp*., it is unlikely that we did not detect IgG antibodies in these species due to non-reactivity of the anti-mouse IgG secondary antibody. Rather, it is more plausible that 1) too few *Praomys sp*. and *Hylomyscus sp*. were tested to detect possible LASV exposure in non-reservoir species or 2) LASV has distinct ecological niches that restricts both competent host replication and interactions between primary LASV reservoirs and other rodent species.

In *Mastomys sp*., mean NP IgG concentrations across all specimens were higher than the mean GP IgG concentrations across all specimens (Figure 2). Although the mean NP IgG concentrations were lower than the mean GP IgG concentrations for *Rattus sp*. specimens, the difference was not statistically significant. The difference in mean IgG concentration in *Rattus sp*. specimens may be skewed by the small sample size of *Rattus sp*. specimens that tested positive for IgG. The observation that more *Rattus sp*. had more specimens test positive for GP IgG than NP IgG (Table 2) was unexpected given that NP is more highly expressed than GP in LASV virions.^15^ Although there have not been documented studies of IgG responses in rats specifically infected with LASV or other mammarenavirues, one study performed intraperitoneal infection of C57/BL6 mice, hamsters, mastomys, and gerbils with similar concentrations of LCMV-Armstrong found vastly different NP IgG ELISA responses between the different rodent species.^28^ Taken together, the results provide further evidence that *Rattus sp*. likely contract and clear LASV infection in an epizootic transmission event, rather than acting as a host reservoir capable of maintaining the virus. Further evaluation of the *Rattus sp*. antibody response to LASV and/or other mammarenaviruses in a controlled laboratory setting are needed to further explore these observations.

A linkage between viral antigen presence and IgG presence is expected in a reservoir host if there is a mechanism of persistent infection which affords the opportunity for sustained viral replication. During a persistent LCMV infection low level viremia (and thus antigen) is maintained despite early and consistent generation of non-neutralizing antibodies.^29^ In addition, viral antigen may also be present towards the end of or after an acute viral infection IgG antibodies begin to be produced. To that end, we observed a strong link between simultaneous antigen and IgG presence in *Mastomys sp*. specimens compared to only antigen presence or only IgG presence; *Rattus sp*. specimens were either only antigen positive or only IgG positive (Table 3, Table S5). However, antigen levels and IgG concentrations were not correlated in Ag+/IgG+ *Mastomys sp*. specimens (Figure 3). Our results contrast with the findings of Demby et. al in Guinea, which demonstrated that antigen and antibody presence in *Mastomys sp*. were mutually exclusive.^5^ Differences in the sensitivity and specificity of the assays deployed may account for the different results obtained. Nonetheless, the strong link between simultaneous antigen and IgG presence in *Mastomys sp*. indicate at least some chronic infections within the primary host reservoir, while the lack of simultaneous antigen and IgG presence in *Rattus sp*. most likely indicates an epizootic transmission event from another infected animal. An important caveat to this study is that specimens were sampled at a single timepoint; it is not possible to exactly determine the stage of infection. The high number of Ag+/IgG-*Mastomys sp*. are likely in varying stages pre- and post-infection, from antigen production before viremia to recovering post-infection prior to the detection of IgG antibodies. Although the Zalgen Pan-LASV RDT shows minimal cross-reactivity to other recombinant mammaeraviruse antigens (L. Branco, personal communication), the IgG ELISAs have not been validated against positive human controls with previous infections to other mammarenaviruses. Thus, it is plausible that other related mammarenaviruses may produce cross-reactivity to the IgG ELISAs and produce false positives.

Here, we have performed antigen RDTs on rodent serum in the field, as well as developed a semi-quantitative ELISA assay to assess the prevalence and strength of IgG in wild-caught rodents in Sierra Leone. This allowed us to contribute zoological surveillance data of LASV prevalence in Eastern Sierra Leone. Although observations such as seasonality and changes in species abundance over time cannot be discerned from our study, these data can be used in conjunction with future longitudinal studies of LASV prevalence. We also evaluated genus level IgG responses in *Mastomys sp*. and *Rattus sp*. to both GP and NP, which has not been previously demonstrated. Evaluating genus-level responses to LASV IgG is important as *Rattus* and *Mastomys* species account for nearly 95% of peri-domestic rodents collected in Sierra Leone and can co-habitat together in homes.^30^ Our ELISA assays, as well as those developed by Soubrier and colleagues hold promise for laboratory studies of mice and rats infected with LASV to further study the natural history and course of infection in primary and secondary hosts.^13^ Furthermore, the ability to screen rodent specimens for potential LASV presence in the field with antigen RDTs, and distinguishing the IgG antibody response to different LASV proteins outside of a BSL-4 laboratory will be useful not only for LASV surveillance in rodents but also to prioritize specimens for downstream analyses to advance Lassa fever knowledge and therapies.

## Data Availability

All data and code are available online at github.com/allisonrs/Project_LASV_AgAb_Rodent

## Notes

## Acknowledgments

We thank Tim Bailey (Bailey and Bailey Reptiles, Folsom, LA, USA) and Heinz Feldman (Rocky Mountain Laboratories, National Institute of Allergy and Infectious Disease, Hamilton, MT, USA), for providing *Mastomys sp*. specimens for control testing; Georgina Dobek and the veterinary staff at Tulane University School of Medicine for assisting with animal handling procedures; and Christopher Bishop and Tynette Hills from Tulane University School of Medicine, Simbirie Jalloh from Kenema Government Hospital, Michelle McGraw from The Scripps Research Institute, and Douglass Simpson from Zalgen Labs for administrative and logistic assistance.

## Contributions

Conceptualization: ARS, JK, JSS, RFG. Field data collection and curation: JK, FK, MF, LK, FA, IMK, MSY, DSG. Laboratory data collection and curation: ARS, MLB, MM, JDS, DJB, DKSN, KMC, RK, MP. Data analysis: ARS, MLB, NGB, JGS. Writing - original draft: ARS. Writing - review and editing: RWC, JSS, RFG. Funding acquisition: KGA, RWC, JSS, RFG. Supervision: JSS, RFG.

## Research in Context

### Evidence before this study

A literature review of previous studies was conducted in PubMed (search terms include “mastomys natalensis lassa”, “lassa serology”, “lassa IgG”, “lassa”, “lcmv serology”, “lcmv IgG”, and “lcmv”) to include studies published prior to September 2022. This was used to determine the number of LASV rodent serology studies conducted in West Africa, diagnostics available for LASV, and potential diagnostic and mechanistic studies performed with LCMV, a surrogate mammarenavirus. There have only been two LASV rodent serology studies in Sierra Leone despite the high burden of LASV and LF in the country. With respect to LASV diagnostics, only one other study specifically developed assays for use in rodent serology studies.

### Added value of this study

Nearly all rodent LASV surveillance studies use indirect immunofluorescence (IFA) to assess for presence of IgG. It is a qualitative method that only assesses presence of LASV rather than the strength of the IgG response. IFA requires advanced biocontainment facilities that are unavailable in West Africa, dramatically increasing costs and resources required to perform the study. Methods to detect relative strength of LASV antigen and IgG as described in this manuscript were performed entirely in-country, allowing rapid identification of current or previous LASV infection in rodents. Furthermore, we use the tools presented in this study to contribute much needed LASV zoonotic surveillance data in Eastern Sierra Leone.

### Implications of all the available evidence

Rapid identification of rodents currently or previously infected with LASV can provide enormous benefit to public health officials in the contexts of outbreak investigation, routine surveillance, and intervention implementation. These tools can also be used to prioritize specimens testing positive for LASV antigen and/or IgG for downstream analyses, enabling further scientific discoveries on the intricacies of LASV in the rodent host reservoir.

