## Supplemental Information for "Novel Tools for Lassa Virus Surveillance in Peri-domestic Rodents"

### Materials and Methods

*Generation of control serum*

Five BALB/C mice (Prosci) and two Sprague-Dawley rats (Thermofisher) were immunized with antigens to either LASV GP (Lineage IV Lassa rGP, Zalgen Labs) or NP (A mixture of Lineage II, II, and IV Lassa N-terminal (1-340) rNP, Zalgen Labs) with Complete Freund's Adjuvant and boosted on days 21 and 35 with Incomplete Freund's Adjuvant. Terminal bleeds were performed on day 50. Equal amounts of terminal bleed from each mouse were mixed to create the mouse ELISA positive control, and equal amounts of terminal bleed from the two rats were mixed to create the rat ELISA positive control. Mouse GP serum was diluted 1:25 in PBS and Mouse NP serum was diluted 1:35 in PBS prior to the 1:100 dilution on the test plate. The initial concentration of the plated 1:100 diluted positive controls were set at 1000 arbitrary units (U)/mL.

Pooled serum from sham-inoculated BALB/C mice (*Mus musculus*) and Sprague-Dawley rats (*Rattus rattus*) was used as ELISA negative controls on their respective experimental plates. *Mastomys sp*., identified by cytochrome *b* PCR as *M. coucha*[1] were obtained from a local python breeder (Bailey & Bailey Reptiles, LA, USA) to initially compare *Mastomys sp*. serum to the sham-inoculated serum (n=40) for establishing negative cutoffs. Additional *M. natalensis* serum (N = 2) was kindly provided by Heinz Feldman of Rocky Mountain Laboratories (NIAID, Montana, USA) to compare with *M. coucha* bleeds. Serum from individual Sprague- Dawley rats (N =20) was purchased (Biomed Chemical, VA, USA) to compare against the sham-inoculated rat serum.

*ELISA Protocol*

Serum from individual specimens and negative controls was diluted 1:100 in sample diluent. A three-fold, six-point standard curve with an initial 1:100 dilution was created from the pre-diluted positive control serum in sample diluent. 100 µL of diluted serum/controls were placed on the antigen coated ELISA plate and incubated at room temperature for 30 minutes. The plate was washed four times x 300 µL/well with PBS-Tween wash buffer using an automatic plate washer. 100 µL/well of the mouse or rat HRP antibody solution were placed on the wells and incubated at room temperature for 30 minutes. The 4 x 300 µL wash step was repeated. 100 µL/well of TMB substrate was added to the plate and incubated at room temperature for ten minutes while protected from light. Finally, 100 µL/well of stop solution was added and plate absorbance was read at 450nm.

*ELISA cutoffs*

To interpolate relative sample concentrations from the standard curve, a four-parameter (4PL) logistic regression model was initially created in GraphPad Prism software v.9.0 (Graphpad, CA, USA) from the optical density (OD) values of all samples and controls on each plate as previously described.[2,3] The initial concentration of the plated 1:100 diluted positive controls were set at 1000 arbitrary units (U)/mL. Due to discrepancies in OD between laboratory and field-collected specimens, the number of positive control animals available, and laboratory temperature and humidity differences between the United States and KGH, receiver operator characteristic curves of values derived from 4PL to create an established cutoff were unable to be used. Instead, the negative cutoff was set at twice the mean of the average negative sample run across the experimental plates. Cutoffs for the IgG ELISAs were as follows: Mouse GP: 28.2 U/mL (N = 6); Mouse NP: 45.9 U/mL (N = 6); Rat GP: 5.24 U/mL (N = 4); Rat NP: 1.41 U/mL (N = 4).

*Statastical analysis*

All statistical comparisons and figures generated for this publication were performed in R (v4.1.3) and RStudio (v1.4.1717) with the packages tidyverse, sf, janitor, ggpubr, ggprism, rstatix, and tayloRswift.[4-11] Shape files of administrative boundaries of Sierra Leone were obtained from the Humanitarian Data Exchange (Sierra Leone -

Subnational Administrative Boundaries - OCHA West and Central Africa- https://data.humdata.org/dataset/cod-ab-sle).

**Results**

**Table S1**: Geographic Positioning System (GPS) coordinates of villages trapped as part of study. Coordinates are given in decimal degrees.

| **Coordinates of Study Villages** | | |
|---|---|---|
| **Village** | **Latitude** | **Longitude** |
| Bambawo | 8.00928 | -11.1325 |
| Bowohun | 8.10533 | -11.2127 |
| Gouma | 8.2869 | -11.0665 |
| Jamboma | 8.121517 | -11.2149 |
| Joru | 7.69242 | -11.0563 |
| Kamboma | 8.128567 | -11.1136 |
| Koi | 8.0412 | -11.0551 |
| Kormolu | 7.995233 | -11.0835 |
| Koyama Ngeima | 8.23473 | -11.1818 |
| Kpalu I | 7.907617 | -11.0849 |
| Kptema | 8.27375 | -11.0744 |
| Largo | 8.05131 | -11.1053 |
| Maleh | 7.898208 | -11.0462 |
| Mano-Ngeiya | 8.0876 | -11.0994 |
| Ngeihun | 8.1748 | -11.0853 |
| Njagor | 8.24533 | -11.1354 |
| Nyahahun | 7.989117 | -11.0849 |
| Pandembu | 8.214267 | -11.066 |
| Panguma (vaama) | 8.186067 | -11.129 |
| Pujehun | 8.15158 | -11.092 |
| Saahun | 7.769487 | -11.1131 |
| Saleima | 8.122449 | -11.1936 |
| Tongola | 8.211017 | -11.0509 |

**Table S2**: Counts of species identification by Cytochrome *b* PCR for tested specimens.

| **Species ID by *Cytb* PCR** | |
|---|---|
| **Species** | **N** |
| N/A: Not tested | 168 |
| *Mastomys natalensis* | 136 |
| *Rattus rattus* | 60 |
| *Praomys rostratus* | 8 |
| *Hylomyscus simus* | 1 |

**Table S3**: Counts of specimens to the genus level as determined with in-field morphometric features, and the type of secondary antibody used for IgG ELISA. *Later identified by cytochrome *b* PCR as *Praomys rostratus.*

| ELISA Secondary Antibody Selection | | |
|---|---|---|
| **Field ID Genus** | **Secondary Antibody** | **N** |
| *Mastomys* | Mouse | 219 |
| *Rattus* | Rat | 139 |
| *Praomys* | Mouse | 11 |
| *Hylomyscus* | Mouse | 3 |
| *Malacomys** | Mouse | 1 |

**Table S4**: NP IgG ELISA results by genus with row percentages. *Includes 1 field identified *Malacomys sp.* specimen later confirmed to be *P. rostarus* by cytochrome b PCR.

| NP IgG Positive Specimens by Genus | | |
|---|---|---|
| **Genus** | **N** | **NP IgG + (% of Genus)** |
| *Mastomys sp.* | 219 | 36 (16) |
| *Rattus sp.* | 139 | 4 (3) |
| *Praomys sp.** | 12 | 0 (0) |
| *Hylomyscus sp.* | 3 | 0 (0) |
| Total | 373 | 40 (11) |

**Table S5**: Contingency table of antigen RDT and IgG ELISA results for *Rattus sp.*†p-values from Fisher's exact test for comparing proportions of antigen (Ag) positive and IgG antibody positive specimens.

| Contingency Table of Antigen and Antibody Presence for *Rattus sp.* | | | | |
|---|---|---|---|---|
| | **IgG -** | **IgG +** | **Total** | **p-value†** |
| **Ag -** | 126 | 10 | 136 | 1 |
| **Ag +** | 3 | 0 | 3 | |
| **Total** | 129 | 10 | 139 | |

**Figure S1**: Correlation between GP IgG and NP IgG concentration from *Mastomys sp.* specimens positive for both GP IgG and NP IgG (N = 22) (Linear regression + standard error).

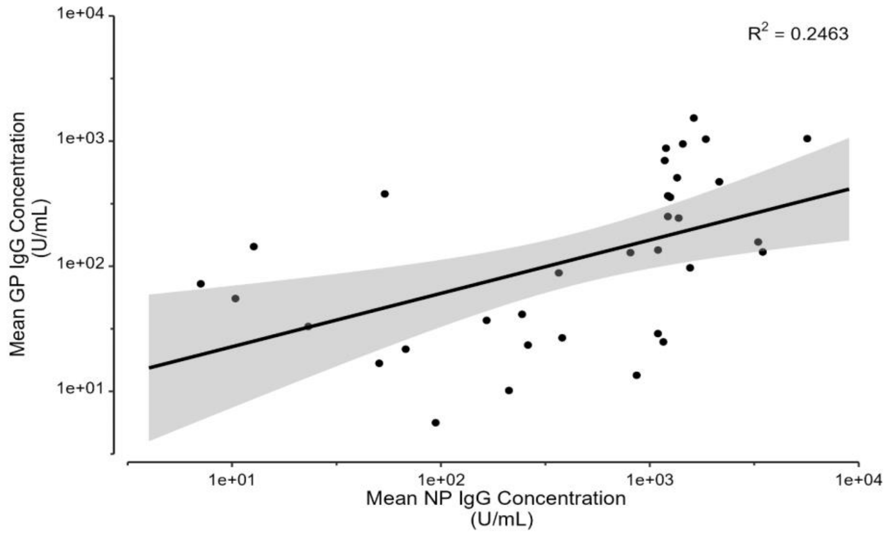
